# Combining antibody markers for serosurveillance of SARS-CoV-2 to estimate seroprevalence and time-since-infection

**DOI:** 10.1101/2021.09.06.21261175

**Authors:** Md S. Bhuiyan, Ben J. Brintz, Alana L. Whitcombe, Alena J. Markmann, Luther A. Bartelt, Nicole J. Moreland, Andrew S. Azman, Daniel T. Leung

## Abstract

Serosurveillance is an important epidemiologic tool for SARS-CoV-2, used to estimate burden of disease and degree of population immunity. Which antibody biomarker, and the optimal number of biomarkers, has not been well-established, especially with the emerging rollout of vaccines globally. Here, we used random forest models to demonstrate that a single spike or receptor-binding domain (RBD) antibody was adequate for classifying prior infection, while a combination of two antibody biomarkers performed better than any single marker for estimating time-since-infection. Nucleocapsid antibodies performed worse than spike or RBD antibodies for classification, but is of utility for estimating time-since-infection, and in distinguishing infection-induced from vaccine-induced responses. Our analysis has the potential to inform the design of serosurveys for SARS-CoV-2, including decisions regarding number of antibody biomarkers measured.

## Introduction

Increasingly, cross-sectional serosurveillance is being used to estimate the fraction of the population previously infected with SARS-CoV-2. Representative seroepidemiological studies reveal the immune landscape of the population, and compared to the use of data based on viral detection, they can provide more accurate insights into the infection fatality rate, the amplitude of transmission in different populations, and highlight disparities in infection rates without typical health-seeking behavior biases [1]. Further, such population-level surveys, when coupled with statistical and/or mechanistic models, could be used to estimate the probability and timing of future waves of disease, measure the impact of interventions such as physical distancing or vaccination, and in later stages, confirm the absence of transmission [2]. However, current knowledge of the kinetics of antibody responses to SARS-CoV-2 infection is insufficient to fully realize the array of use-cases for data from population-level seroepidemiological studies. For those designing serosurveys, the choice of antibody assays can be daunting given the number of available tests that target different antigens and isotypes. The aim of this study was to provide new evidence to highlight the best antibody biomarkers for estimation of seroprevalence and time-since-SARS-CoV-2 infection, and whether a combination of antibody biomarkers could improve such estimations.

## Methods

### Data sources

We identified studies in the literature or on preprint servers that measured multiple antibody responses at varying time points greater than a median of 50 days after PCR-confirmed SARS-CoV-2 infection [3-7]. Data that were not publicly available were obtained on request from study authors. Antibody responses examined included IgG, IgM and IgA responses against spike (S), receptor binding domain (R) and nucleocapsid (N) antigens as determined by ELISA or Luminex multiplex bead assays. For each serologic measurement, the time between serologic sample collection was extracted and either the day of PCR confirmation or symptom-onset, which was termed “time since infection.” For subjects with antibody response measurements at more than one time point, only the last time point was used. This analysis was reviewed by the Institutional Review Board of the University of Utah and determined to not meet definitions of Human Subjects Research.

### Outcomes and predictor variables

We explored how individual, and combinations of, antibody measurements could identify those who were infected with SARS-CoV-2 and, if infected, their time since last infection. Using antibody biomarkers measured at different time points post-infection and those collected before the SARS-CoV-2 pandemic, we evaluated the importance and performance of IgG, IgM and IgA antibody isotypes against the nucleocapsid (N), the spike surface protein (S) and receptor binding domain (R) antigens in 1) identifying previously infected individuals, and 2) their time since infection. We only used binding antibody biomarkers and excluded neutralizing antibody results due to the complexity of the assay and variability in methodology.

### Model development

We used random forest models to both determine the order of importance of biomarkers and to make our final predictions (1000 random trees, 3 biomarkers per split). Due to the highly correlated nature of the biomarkers, we measured importance using the conditional permutation importance algorithm [8], which measures the importance of each biomarker conditioned on other associated biomarkers in the model. We used this order of importance to train and test various sized sets of the antibody response predictors. We fit the models for each study separately, using the cforest and party packages, and measured importance using the permimp package, in R [8, 9].

### Model comparison

In order to assess the predictive performance of each model, we used repeated 5-fold cross-validation (CV) with 100 iterations, where each iteration contained screening (variable importance) and model-fitting steps. Within the cross-validation, we considered predictor sets of sizes 1, 2, or 3 variables. We developed reduced compact models with a maximum of three predictor antibody responses to make our models more applicable to public health practice. We also investigated performance of the full model in all datasets with available biomarker variables to understand the upper limit. Using this process, we developed two independent models: 1) a model to identify PCR-confirmed infections using biomarkers and 2) a model to estimate time since infection among those previously infected. For the first model, we used PCR confirmed cases and pre-pandemic controls to train a random forest model and assessed performance using the cross-validated area under the curve (cvAUC) [10]. Within each iteration of cross-validation, we compared model ROC curves using a permutation test with the function roc.test. We then summarized the p-values across iterations to compare model performance [11]. For the second model, we used only confirmed cases to train a random forest model for estimating time-since infection and assessed performance using cross-validated mean absolute error (MAE), the mean of the absolute differences of the predictions from the true time since infection.

## Results

We extracted and analyzed data on PCR-confirmed SARS-CoV-2 infections from 5 separate studies, with a total of 834 subjects (Table 1). Of the 5 studies, 4 of them used enzyme immunoassays, while 3 of them used Luminex bead array methods. Among a total of 834 subjects, the median time from infection to serologic sampling was 76 [IQR 51-98] days, the median age was 49 [IQR 33-60] years, and the proportion of males was 46.4 percent. Among the 5 studies, the proportion of patients with severe disease or those who were hospitalized ranged from 7 to 24 percent.

**Table 1.** Summary of the characteristics of datasets used in this analysis, with cross-validated AUC (95% CI) from classifying previous infection on four of the published datasets. The rows with row-name starting “Best” include a screening step in which the biomarkers are ordered by importance for classification (ever-infected) using the random forest conditional permutation algorithm and only the top biomarkers from that iteration are used when training the model.

|  | Dan et. al.<br>[3] | Peluso et.<br>al. [6] | Whitcomb<br>e et. al. [7] | Markmann<br>et. al. [5] | Isho et. al.<br>[4] |
| --- | --- | --- | --- | --- | --- |
| No. of patients | 101 | 122 | 112 | 156 | 343 |
| Age, Median (IQR) | 38 (29.5-46.5) | 48 (19-85) | 48(31-57) | 46.5 (32-60) | 36 (21-44) |
| Sex (Male, %) | 45 | 54.9 | 43.7 | 44.0 | 52.1 |
| % patients with severe disease (hospitalized) | 7 | 24.2 | 13.7 | 17.7 | n/a |
| Median time since infection (IQR) | 62 (47.5-87) | 115 (95-124) | 109 (94.7-190.5) | 57 (49-79) | 72 (51-86) |
| Immunoassay platform | ELISA | ELISA/Luminex | Luminex | ELISA/Luminex | ELISA |
| Cross validated AUC (95% CI) |  |  |  |  |  |
| RBD IgG | n/a | 97.8 (97.2-98.5) | 99.7 (99.4-99.9) | 98.7 (97.9-99.5) | 96.9 (96.6-97.3) |
| Spike IgG | n/a | 98.8 (98.3-99.2) | 99.7 (99.5-99.9) | n/a | 99.1 (98.8-99.3) |
| Nucleocapsid IgG | n/a | 97.9 (97.4-98.4) | 93.9 (93.2-94.5) | 98.9 (98.2-99.5) | 94.4 (93.9-94.9) |
| Nucleocapsid IgG vs Best of Spike/RBD IgG.<br>Mean (SD) p-value, proportion of iterations | n/a | 0.27(0.27),<br>0.24 | 0.38(0.28),<br>0.11 | 0.36(0.30),<br>0.15 | 0.40(0.28),<br>0.1 |
| with p-value <0.05 |  |  |  |  |  |
| Best Two biomarkers | n/a | 98.8 (98.4-99.2) | 99.9 (99.9-99.9) | 99.2(98.6-99.8) | 99.4 (99.2-99.5) |
| Spike/RBD IgG vs Best Two biomarkers.<br>Mean (SD) p-value, proportion of iterations with p-value <0.05 |  | 0.62(0.31),<br>0 | 0.28(0.28),<br>0.22 | 0.23(0.29),<br>0.28 | 0.19(0.26),<br>0.48 |
| Best Three biomarkers | n/a | 99.0 (98.7-99.4) | 99.9 (99.9-99.9) | n/a | 99.2 (99.1-99.4) |
| Best Two vs Best Three biomarkers.<br>Mean (SD) p-value, proportion of iterations with p-value <0.05 | n/a | 0.30(0.31),<br>0.11 | 0.35(0.29),<br>0.14 | n/a | 0.16(0.25),<br>0.48 |
| Best Two Nucleocapsid biomarkers | n/a | n/a | 95.0 (94.5-95.5) | n/a | 95.9 (95.5-96.2) |
| Full/saturated model | n/a | 99.0 (98.8-99.3) | 99.9 (99.9-99.9) | n/a | 99.4 (99.3-99.5) |

### A single antibody biomarker sufficiently identifies previous infection

We first explored the classification performance of single antibody and isotype thresholds in identifying infection using 4 of the 5 datasets with pre-pandemic control data available. We show that across all studies, a single RBD or spike IgG biomarker performs similarly to the combination of the best two biomarkers in identifying prior infection (Table 1). Addition of a third biomarker did not increase discriminatory performance in any of the studies examined and further addition of biomarkers resulted in no additional performance benefit (p > 0.05 for all studies).

### Two antibody biomarkers are better than one for prediction of time-since-infection

Next, we explored the performance of single vs multiple biomarker thresholds in predicting time-since-infection. In all 5 datasets, combining two antibody biomarkers performed better than the best single IgG for estimation of time-since-infection (Table 2). Using conditional permutation variable importance from random forest regression to measure the mean decrease in accuracy importance, we found that in the 4 datasets where multiple antibody isotypes are measured, the best two antibody biomarkers included a combination of an IgG and an IgM (or IgA in the one dataset where IgM was not measured, Figure 1). Addition of a third marker results in a marginal (within SD) increase in prediction performance in 3 of the 5 datasets (Table 2).

**Table 2.** Mean (standard deviation) of MAE from predicting time since infection from repeated cross-validation on five published datasets. The rows with row-name starting “Best” include a screening step in which the biomarkers are ordered by importance for time-since-infection using the random forest conditional permutation algorithm and only the top biomarkers from that iteration are used when training the model (low MAE indicates better performance).

|  | Dan et. al.<br>[3] | Peluso et.<br>al. [6] | Whitcombe<br>et. al. [7] | Isho et. al [4] | Markmann et.<br>al. [5] |
| --- | --- | --- | --- | --- | --- |
| Best RBD IgG | 18.7(1.7) | 29.2(3.7) | 59.8(5.4) | 17.3(1.1) | 21.4(5.7) |
| Best Spike IgG | 19.6(1.9) | 26.6(3.6) | 61.1(6.9) | 17.5(1.3) | n/a |
| Nucleocapsid IgG | 18.8(2.0) | 23.9(4.0) | 55.9(7.4) | 18.3(1.4) | 21.2(6.3) |
| Best Two biomarkers | 17.1(1.9) | 22.6(4.4) | 53.1(5.1) | 15.7(1.4) | 20.9(5.3) |
| Best Three<br>biomarkers | 17.1(1.9) | 22.4(4.0) | 52.5(5.3) | 15.3(1.4) | n/a |
| Best two<br>Nucleocapsid<br>biomarkers | n/a | 24.7(3.9) | 51.3(7.1) | 17.5(1.1) | n/a |
| Full/saturated model | 17.8(1.9) | 22.2(3.8) | 51.5(6.3) | 15.1(1.1) | 20.6(5.9) |

**Figure 1.**
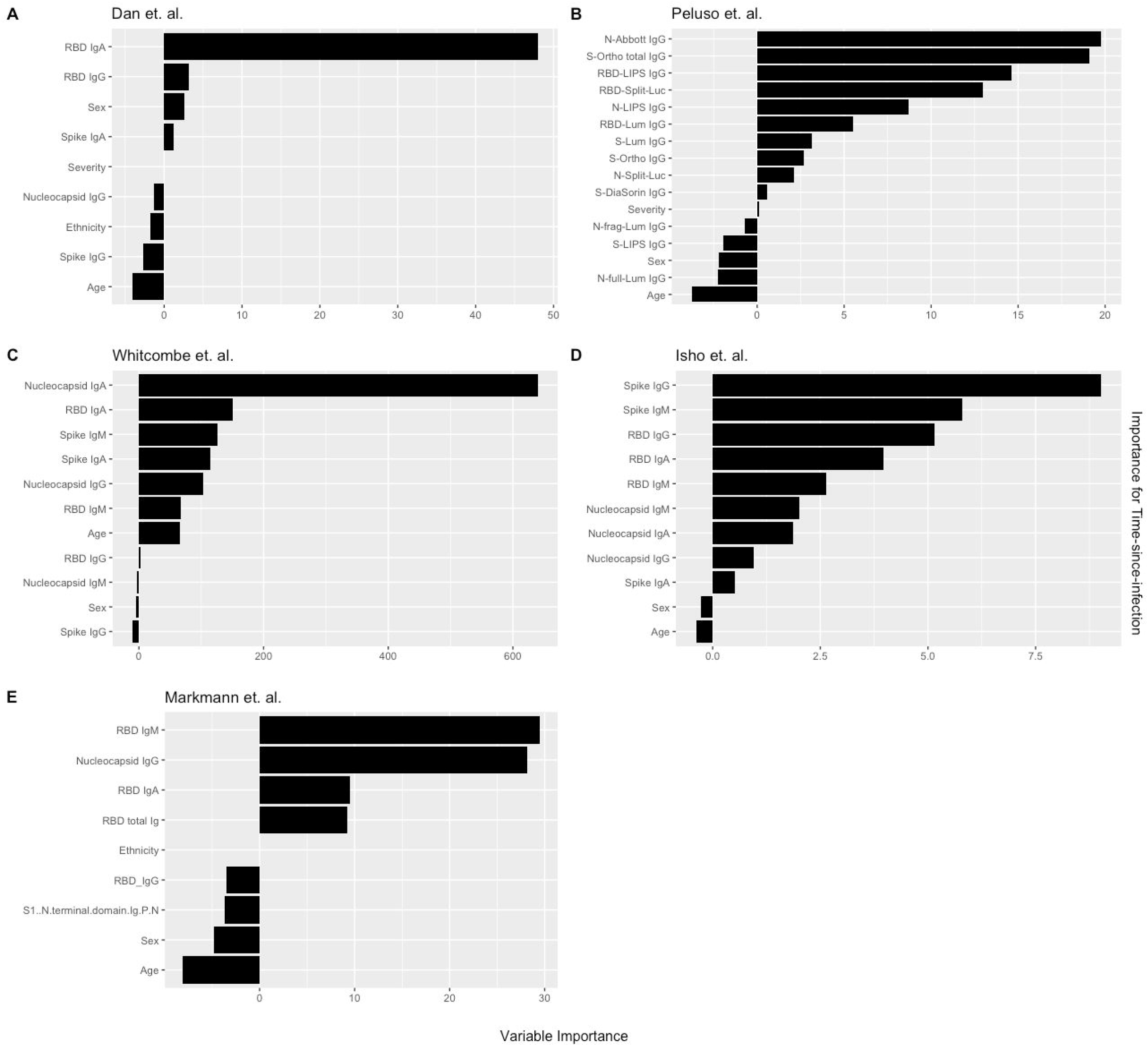
Conditional permutation variable importance from random forest regression measured by mean decrease in accuracy. Negative importance indicates that the variables inclusion has decreased mean accuracy, probably due to overfitting or random error. Each column represents the order of importance of biomarkers in five datasets. Abbreviations: In Peluso et al dataset, S_Ortho_Ig and S_Ortho_IgG indicate total Ig and S IgG by Ortho Clinical Diagnostics VITROS kits; N_abbott indicate Abbot ARCHITECT (IgG); S_DiaSorin is Spike IgG by DiaSorin LIASON(IgG); Neu_Monogram is Monogram PhenoSense (neutralizing antibodies); RBD_LIPS, S_LIPS, N_LIPS is IgG by Luciferase Immunoprecipitation System (LIPS); RBD_Split_Luc, N_Split_Lum, S_Lum, N.full_Lum, N.frag_Lum indicate IgG to respective antigens by Luminex assay.

### Nucleocapsid antibody biomarkers are suboptimal for classification of previous infection, but adequate for estimating time-since-infection

Given that all vaccines currently approved for use in USA/EU induce spike or RBD antibody responses, we examined the performance of nucleocapsid-only combinations of antibodies. For identification of previous infection, nucleocapsid IgG performed statistically significantly worse than RBD/spike IgG in 2 of 4 studies examined. In the two studies where data were available, the combination of the two top nucleocapsid markers (IgG plus either IgM or IgA) improved discriminatory performance (Table 1). On the other hand, for predicting time-since-infection (Figure 1), a combination of the two top nucleocapsid markers performed similar to, or better than, RBD or spike IgG alone (Table 2).

## Discussion

The current COVID-19 pandemic is a major public health concern worldwide, and assessment of infection burden in populations is crucial towards efforts to mitigate its spread and inform policy and decision-making. Population-level serosurveillance has emerged to be a useful method to provide accurate estimates of disease burden, as when done under a representative sampling framework, is not subject to biases related to health-seeking behavior or testing availability. However, there are limited studies to inform the choice and numbers of antibody biomarkers for SARS-CoV-2 serosurveillance. Here, we built models using serologic data from five studies of individuals with confirmed SARS-CoV-2 infection, to examine which biomarker(s) are best for identifying prior infection and prediction of time-since-infection. Our results show that while Spike/RBD IgG alone are adequate for discrimination/classification of those who have been infected, combinations of antibody markers may be best for estimation of time-since-infection.

An important consideration in the design of serosurveys is the selection of the biomarker(s), with a goal of minimizing cost while capturing enough information about infection, transmission or immunity. Population-level serosurveys are able to not only provide estimates for seroprevalence (proportion who have been infected), they also have the potential to provide data towards estimating the time-since-infection, which could help with accurate incidence estimation and tracking transmission changes on a population level. Our analysis, using mean absolute error as a performance measure, suggests that a combination of antibodies are the best predictors of time-since-infection. For the majority of studies examined, we found using that using 3 or more biomarkers only performed slightly better than use of only two biomarkers. In addition, we show that clinical-demographic factors such as age (and less so severity) were potentially important predictors that should be considered in model-building. Further studies are needed to assess the combined performance of both stages of this model by recreating the epidemic curve through estimation and comparing it to a known epidemic curve.

As COVID-19 vaccines are increasingly made available worldwide, distinction of vaccine-induced immune responses from that elicited by natural infection is important in the design of seroepidemiologic studies. Identifying infections in vaccinated populations will help estimate the rate of spread. Unfortunately, the most widely-used antibody markers for SARS-CoV-2 serosurveillance are the IgG to S or RBD, which is also the target of all currently approved vaccines in the US/EU. Thus, future serosurveillance efforts may increasingly depend on the nucleocapsid antibody. Our analysis suggests that while nucleocapsid specific IgG alone is inferior to spike and RBD for classification of infection, combinations of N antibodies may improve performance. Notably, for estimation of time-since-infection, the best two N antibodies performed similarly or better than any single S or RBD antibody. Thus, monitoring nucleocapsid specific antibodies may be of utility in distinguishing infection-related antibodies from vaccine induced antibody response.

A number of research questions and goals remain for SARS-CoV-2 seroepidemiology. First, our conclusions regarding use of biomarkers for SARS-CoV-2 serosurveillance are based on internal cross-validation of models built using datasets featuring antibody responses for up to 200 days from time of symptoms onset or diagnosis. Detailed characterization of the kinetics of serologic responses through longitudinal cohort studies of infected persons of varying severity will enable development of more tailored and precise statistical models of recent infection. Second, in addition to commercial platforms, standardization of serosurvey reagents, such as through publicly-available monoclonal antibody standards, and/or reference positive sera, will enable a broader application and validation of seroepidemiological analytical models. Third, development of point-of-care antibody testing will enable serosurveillance to be better performed in more austere environments. Use of dried blood spots from finger pricks in low resources settings could reduce cost while obviating need for cold-chain storage [7]. Fourth, high-throughput multiplex platforms such as Luminex technology [12] could enable the measurement of numerous SARS-CoV-2 serological markers alongside markers against other infectious pathogens of interest.

There were a number of limitations in this analysis. The lack of longitudinal immune responses and lack of detailed time-since-infection data that may have led to larger error predicting time-since-infection. Our analysis was limited to studies of adults in high-income countries, and thus our results cannot be generalized to low- and middle-income countries, or to pediatric populations, and underscore the need for a better understanding of the kinetics of SARS-CoV-2 antibody responses across diverse populations. Despite this, our findings contribute towards informing the choice of antibody responses for seroepidemiological investigations of SARS-CoV-2.

## Data Availability

All code and data will be deposited on github upon publication.

## Notes

### Author contributions

M.S.B., A.S.A., and D.T.L. designed and directed the project. M.S.B. and B.J.B. contributed to data analysis. M.S.B. and D.T.L. wrote the paper. All authors discussed the results and commented on the manuscript.

### Financial support

This work was supported in part by the National Institutes of Health (R01 AI135115 to D.T.L.), with funding in part from the National Center for Research Resources and the National Center for Advancing Translational Sciences of the National Institutes of Health, through Grant UL1TR002538 (formerly 5UL1TR001067-05, 8UL1TR000105 and UL1RR025764).

### Potential conflicts of interest

All authors reported no conflicts of interest.

### Data Availability

All code and data will be deposited on github upon publication.

## Notes

### Competing Interest Statement

The authors have declared no competing interest.

### Author Declarations

This analysis was reviewed by the Institutional Review Board of the University of Utah and determined to not meet definitions of Human Subjects Research.

